# Intravenous streptokinase for myocardial infarction: a reanalysis of the historical cumulative evidence using Trial Sequential Analysis

**DOI:** 10.1101/2024.09.24.24314267

**Authors:** Kim Boesen, Christian Gluud

## Abstract

**Introduction:** Cumulative meta-analysis of intravenous streptokinase for myocardial infarction has been used as a text-book example to show how the megatrials GISSI and ISIS-II were redundant and wasteful. We decided to reanalyse the dataset with Trial Sequential Analysis to account for statistical heterogeneity and the risk of bias of the historical trials to reassess whether GISSI and ISIS-II were justified or redundant.

**Methods:** We extracted data from overviews published in 1982 and 1985 and trial reports on mortality outcomes. For the five largest trials conducted before GISSI and ISIS-II, we also extracted information on the used comparator, randomisation, blinding, dropout proportions, and the use of intention-to-treat analyses. We did random-effects cumulative meta-analyses with Trial Sequential Analysis considering diversity.

**Results:** The largest trials conducted before GISSI and ISIS-II had serious methodological differences and high risks of bias making a cumulative meta-analysis invalid by today’s standards of evidence synthesis. The Trial Sequential Analysis showed that the monitoring boundary for a mortality benefit of streptokinase was reached during the ISAM trial. However, both GISSI and ISIS-II were launched before the ISAM trial was published. Focusing only on the cumulative assessment, the megatrials were potentially futile. Sensitivity analyses corroborated these results.

**Conclusion:** Our Trial Sequential Analysis of the historical dataset of streptokinase for myocardial infarction found that conclusive evidence favouring streptokinase was established after the megatrials were launched. However, considering the methodological differences and risks of bias, such cumulative meta-analysis seems invalid. Accordingly, the megatrials were not wasteful.

## Introduction

GISSI (1984 to 1985)^1^ and ISIS-II (1985 to 1987)^2^ were two megatrials assessing intravenous streptokinase (fibrinolytic intervention) for myocardial infarction. GISSI enrolled almost 12,000 participants to streptokinase plus usual care versus usual care only. ISIS-II enrolled more than 17,000 participants in a factorial design to streptokinase versus aspirin versus the two in combination versus placebo for myocardial infarction. GISSI reported an overall mortality risk reduction of 19%, 95% confidence interval (CI) 10% to 28%. ISIS-II found that aspirin and streptokinase alone compared with placebo reduced the odds of 5-week vascular mortality with 23% and 25%, respectively, and the combination of aspirin and streptokinase reduced the odds with 42%. ISIS-II has attained mythical status and is considered one of cardiology’s most influential trials.^3,4^

In 1992, Lau and colleagues casted doubts about whether these megatrials were ethically and scientifically justifiable.^5^ Using a statistical method called cumulative meta-analysis, which is a meta-analysis that is updated after each new trial, they found that a statistically significant reduction in mortality of streptokinase compared with various controls could be demonstrated after eight trials already in 1973, 11 years before GISSI was launched. ^5^ This summary estimate did not change meaningfully with the addition of the subsequent 25 trials.^5^ However, one cannot simply assess the result of a cumulative meta-analysis as conclusive the first time the summary estimate reaches statistical significance. To draw definitive conclusions from a cumulative meta-analysis, one needs to consider (1) the number of randomised participants in relation to the meta-analytic sample size (i.e. number of participants needed to draw definite conclusions); (2) the multiple comparisons problem, or multiplicity (i.e. the risk of random error from continuously making inference testing); (3) whether the trials were at risk of bias; and (4) heterogeneity (i.e. the statistical variation between the trial results).^6-9^ Moreover, as a condition for any meaningful meta-analysis, characteristics such as patient population, administered intervention and control, and outcomes should be comparable across trials.

To address the multiple comparisons problem, Pogue and Yusuf (one of the original ISIS-II investigators) reanalysed the streptokinase meta-analysis in 1997^6, 7^ as if the data came from one big trial. First, they calculated the sample size of patients needed to demonstrate a certain treatment effect and called it the ‘optimal information size’. Secondly, to avoid drawing wrong conclusions before this sample size was reached in the meta-analysis, they introduced monitoring boundaries known from interim analyses in a single trial. In other words, the threshold for considering the meta-analytic result statistically significant depends on the number of analysed participants; the fewer participants, the more extreme the result has to be, to avoid the risk of spurious false positive results. Pogue and Yusuf’s reanalysis did not consider heterogeneity between the included trials neither the risk of bias of the trials, or whether they should be meta-analysed at all. An established method to account for the heterogeneity is random-effects Trial Sequential Analysis.^8, 9^

We decided to update Pogue and Yusuf’s analysis by applying Trial Sequential Analysis on the historical trial data on streptokinase for myocardial infarction. We aimed to empirically estimate if, and when, the evidence was conclusive in favour of streptokinase, and thus whether the megatrials GISSI and ISIS-II should be considered justified or futile.

## Methods

### Data source

We extracted data from two systematic reviews published by the ISIS-II investigators in 1982^10^ and 1985,^11^ to gain information (number of patients and events, i.e. death) from randomised clinical trials conducted before GISSI and ISIS-II comparing intravenous streptokinase with various comparators, i.e. heparin (eight trials), placebo (four), no control (four), glucose (two), albumin (one), and one without information. To extract information on trial designs, we supplemented with data from the European 2,^12^ Australian 1,^13^ UK Collaboration,^14^ N German Collab,^15^ Austrian,^16^ ISAM,^17^ GISSI,^1^ and ISIS-II (only including data from the streptokinase only and the placebo only groups)^2^ trial reports. We did not include a number of smaller trials published between 1986 and 1988 in our analyses.^6^ Data from these small trials was most likely not available to the GISSI and ISIS-II investigators, and their size make them negligible to this reanalysis.

### Risk of bias

We made a simple risk of bias assessments of the five largest trials preceding GISSI plus of the GISSI, ISAM, and ISIS-II trials. We assessed randomisation, blinding, dropout proportions, and the use of intention-to-treat (ITT) analysis.

### Trial Sequential Analysis

We conducted a random-effects model cumulative meta-analysis with inverse variance and DerSimonian-Laird method and the relative risk as a measure for mortality. To run the Trial Sequential Analysis, one needs to determine five parameters: event proportion in the control group (i.e. the proportion of participants dying in the control group); relative risk reduction of the intervention (i.e. the anticipated effect of streptokinase on reducing mortality); alpha (i.e. the threshold for statistical significance); beta (i.e. power to detect a significant difference); and diversity (a measure of the heterogeneity in the meta-analysis).^8, 9^ We used the TSA Software version 0.9.5.10 Beta to run the analyses.^18^ We did not preregister a protocol for this analysis.

We used the all-cause mortality proportion of 19% in the control group in the 1985 review.^11^ We estimated a realistic risk reduction to 20% based on the reported intervention effects in the two systematic reviews. The risk ratio was 0.80 (95% CI 0.68 to 0.95) for mortality favouring streptokinase in 1982 (based on 8 trials),^10^ and the odds ratio on overall mortality was 0.76 (95% CI 0.64 to 0.88) in 1985.^11^ We set alpha to 5% (we only looked at one outcome), and beta to 10% (giving a power of 90%). We used a calculated diversity of 52%, corresponding to the 1985 meta-analysis, to mimic the GISSI and ISIS-II investigators’ contemporary knowledge at their time of decision to launch the trials. In comparison, the GISSI investigators estimated a control event proportion of 10%; mortality risk reduction of 20%; alpha 1%; and power of 95% (beta 5%).^1^ The ISIS-II trial publication did not report sample size calculations.^2^

### Sensitivity analyses

To test the robustness of our analysis, we did two sensitivity analyses with diversity equaling 25% and 75%, a third analysis assuming Pogue and Yusuf’s 1997 parameters (control event proportion 10%, relative risk reduction 15%, alpha 5%, power 90% (beta 10%))^6^ with our calculated diversity of 52%, and a fourth analysis including only trials with a placebo or no intervention control group (according to the 1985 overview), thereby excluding trials with active comparators.

## Results

### Risk of bias

There were serious risks of biases and methodological differences among the eight largest streptokinase trials (Table 1). These include GISSI’s lack of blinding, ISIS-II reporting vascular and not total mortality, the time span of reported mortality from three weeks to 6 months, and the used comparators, such as Europe 2’s comparison of streptokinase with heparin and Australian 1 administering heparin and warfarin to both groups in addition to streptokinase as the intervention. One should challenge the usefulness of meta-analysing such different trials and moreover applying Trial Sequential Analysis on such a collection. But that is what the meta-analyses conducted in the previous assessments until now did.^5, 6, 10, 11^

**Table 1.** Risk of bias in the eight largest streptokinase trials.

| **Trial (size)** | **Study period** | **Comparator** | **Outcome** | **Randomisation** | **Blinding** | **Dropout proportion** | **Intention -to-treat (ITT) analysis** |
| --- | --- | --- | --- | --- | --- | --- | --- |
| European 2 (764)<sup>12</sup> | 1967 –1970 | Heparin (10.000 IU) | Total mortality during hospitalisation (about 6 weeks) | Drawing cards | Not blinded | 4.5% (34 of 764) | Per protocol (removal of one trial site) |
| Australian 1 (517)<sup>13</sup> | 1968 – 1971 | Heparin (5000 IU) | Total mortality at 3 months | Sealed envelopes | Not blinded | Not reported | Per protocol (17 were withdrawn) |
| UK Collab (595)<sup>14</sup> | 1971 – 1974 | No control (usual care) | Total mortality at 6 months<sup>a</sup> | Procedure not described | Not blinded | Not reported | Per protocol (protocol violation of four patients) |
| N German Collab (483)<sup>15</sup> | 1972 – 1974 | No control (usual care)<sup>b</sup> | Total mortality during hospitalisation (about 6 weeks) | Numbered envelopes | Not blinded | Not reported | Not reported |
| Austrian (728)<sup>16</sup> | July 1972 – June 1975 | No control (usual care) | Total mortality at 6 months | Procedure not described | Not blinded | Not reported | Not reported |
| ISAM (1,741)<sup>17</sup> | Mar 1982 – Mar 1985 | Placebo | Total mortality at 3 weeks | Not reported | Blinded | Not reported | Not reported |
| GISSI (11,806)<sup>1</sup> | Feb 1984 – Jun 1985 | No control (usual care) | Total mortality at 3 weeks | Computer generated | Not blinded | 0.9%<sup>c</sup> | Per protocol<sup>c</sup> |
| ISIS-II (17,187)<sup>2</sup> | Mar 1985 – Dec 1987 | Placebo | Vascular mortality at 5 weeks | Computer generated | Blinded | 3% at 5-week follow-up | ITT |

a) Using the same data as Yusuf 1985 review. Mortality also reported at 6 weeks (Table VII).
b) It seems that the intervention group also received heparin and marcumar.
c) “The data sheets of 94 randomised patients (45 SK and 49 C) could not be traced. The baseline characteristics, collected at randomisation, of these "missing" patients (who constituted only 0.9% of the total) closely correspond to those of the sample analysed”.

### Trial Sequential Analysis

Our Trial Sequential Analysis suggests that the cumulative effect estimate for a mortality benefit of streptokinase breached the monitoring boundary already at the end of ISAM in 1985 (Figure 1). This means that the effect was so significant that conclusive evidence was established before the required information size was accrued, which happened during GISSI (Figure 1). The required information size, i.e. the number of participants needed to reach conclusive evidence under the assumptions described above, was 8,597 participants using a diversity of 52%. In summary, ignoring the trials’ differences and risks of bias, GISSI and ISIS-II seem potentially redundant and wasteful according to our analysis and assumptions.

**Figure 1.**
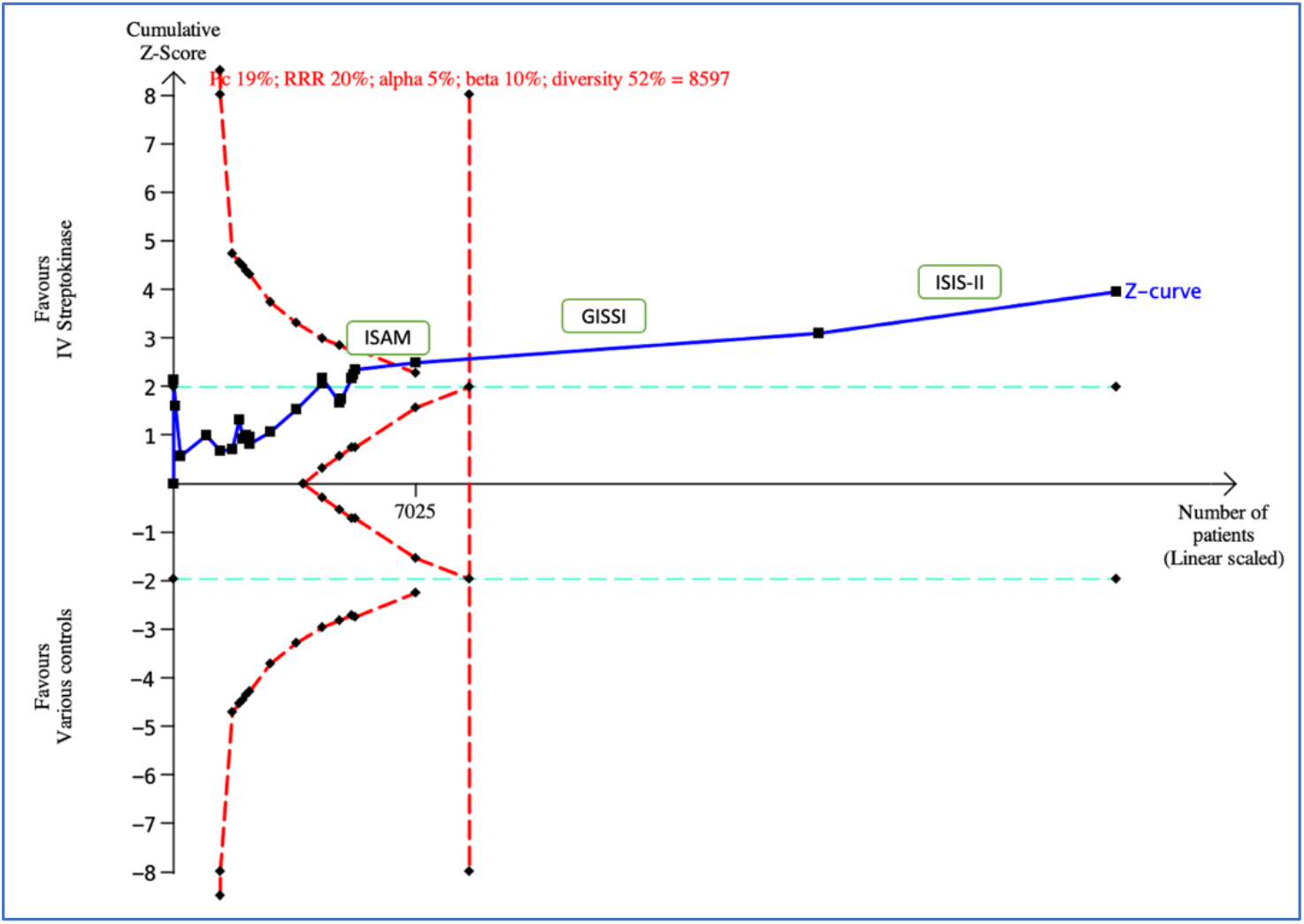
Trial Sequential Analysis of randomised clinical trials comparing streptokinase versus various comparators. How to read the TSA graph: The X-axis shows number of participants included in the trials as well as the required information size of 8,597 participants (the vertical red dotted line). The required information size was calculated based upon the proportion in the control group with the outcome (19%); the relative risk reduction of 20%; alpha of 5%; beta of 10%; and diversity of 52%. The Y-axis shows the Z-score, which is the statistical difference between the two intervention groups, in this case streptokinase versus control. The two horizontal green dotted lines represent -/+ Z=1.96. The two symmetrical inward sloping red dotted lines represent the monitoring boundaries for benefit or harm. The two symmetrical outward sloping red dotted lines represent the monitoring boundaries for futility. Each black dot represents a single trial, and the solid blue line indicates the cumulative Z-score. The ISAM, GISSI, and ISIS-II trials (see text) are shown above their respective Z-scores.

### Sensitivity analyses

Our sensitivity analyses consolidate our main analysis. With diversity of 25%, the required information size was 5,502 participants, and the cumulative summary effect breached both the monitoring boundary for benefit and for futility close to the required information size (Appendix, Figure 1). With diversity of 75%, the required information size increased to 16,508 participants, and it was breached at the end of GISSI, comparable to our main analysis (Appendix, Figure 2). Repeating the Pogue and Yusuf conditions with diversity of 52%, the required information size increased to 32,668 participants but the monitoring boundary for benefit was still breached during GISSI (Appendix, Figure 3). Including only the six placebo-controlled trials and the five trials with no intervention control, the summary effect crossed the boundary for futility during GISSI, but then approached significance at the completion of GISSI (Z=1.96), which was augmented by ISIS-II (Z=3.33) (Appendix, Figure 4).

## Discussion

Our cumulative analyses challenge the importance of the megatrials GISSI and ISIS-II and we estimate that the evidence demonstrating benefit of streptokinase on mortality may have been established by the ISAM trial.^17^ Although our calculated meta-analytic sample size (8,597 participants) differs by a factor 2 to Pogue and Yusuf’s 15,560 participants, our conclusions are comparable; conclusive evidence was likely established during GISSI, and before ISIS-II. The reason for Pogue and Yusuf larger sample size were their more conservative assumptions of a control event rate of 10% and risk reduction of 15%.^6^

However, we find the largest streptokinase trials so clinically and methodologically different that the cumulative meta-analysis does not seem valid. By today’s standards of evidence synthesis such cumulative meta-analysis would not pass as good research to mix completely different comparisons, such as streptokinase versus heparin and streptokinase versus glucose in the same meta-analysis. The trials also had serious risks of bias, including uncertain randomisation procedures and were unblinded, and even reported different outcomes. Before GISSI and ISIS-II, only one large (n=1,747), blinded, and placebo-controlled trial was conducted, the ISAM trial.^17^ This speaks definitively in favour of conducting both GISSI and ISIS-II, although the lack of blinding in the former would not live up to present day scrutiny.^19^

Our present results must also be interpreted in a historical context. Systematic reviews and meta-analyses were at their infancy in the mid 1980s and the medical community was hesitant to accept their results.^4, 20^ In the 1985 review, the ISIS-II investigators stated: “This review of the data clearly indicates that IV fibrinolytic agents can reduce mortality after MI. The effect is highly significant […], substantial […], and reliably estimated […], and cannot be accounted for by any plausible biases”.^11^ Despite this, the investigators felt it was necessary with a large-scale trial to confirm this result.^20^ We agree with this decision, but for the opposite reasons as stated in their 1985 review: there was no reliable and comparable evidence before ISAM and so ISIS-II – and GISSI – were justified.

This reanalysis illustrates the importance of properly assessing the basics of trial designs before meta-analysis. It also illustrates that the higher bar for evidence in your cumulative analysis, the fewer trials you can likely include. This was also the conclusion in an analysis of tranexamic acid to control surgical bleeding.^21^ The cumulative evidence of 128 tranexamic acid trials crossed the monitoring boundary for benefit and the required information size; 38 low risk of bias trials crossed the monitoring boundary but not the required information size; and the analysis with the two prospectively registered trials and prespecified primary outcomes was far from conclusive. As a trialist, one can probably always find reasons to conduct a new trial, and so it becomes a tradeoff between thoroughly assessing existing evidence but not setting the bar so high that – acceptable – conclusive evidence can never be achieved. Finally, we want to highlight the lack of a registered protocol as an important limitation to our reanalysis. The breadth of analytic choices, even in a simple reanalysis of one outcome, open up a multiverse of alternatives, such as inclusion criteria, risk ratio compared to odds ratio, or the DerSiminion-Laird compared to the Sidik Jonkman method.

We encourage researchers to preregister and publish detailed protocols for their systematic reviews and meta-analyses, especially when it comes to using statistical methods, such as the Trial Sequential Analysis.^22^

## Conclusion

Our Trial Sequential Analysis of the historical dataset of streptokinase for myocardial infarction found that conclusive evidence favouring streptokinase was established before the megatrials GISSI and ISIS-II were finished. However, considering the methodological differences and risks of bias, such cumulative meta-analyses seem invalid. Accordingly, the megatrials were not wasteful. We therefore conclude that ISIS-II and GISSI were justified based on the lack of existing evidence from large, blinded, placebo-controlled trials. This reanalysis illustrates the importance of justifying and determining whether a new clinical trial is needed; not just on the totality of evidence and summarised effect estimates, but on the basis of a thorough assessment of preceding trials’ designs and biases. Only when the trials can reliably be meta-analysed, one should use Trial Sequential Analysis to determine if, and when, conclusive evidence has been established, or if further trials are needed.

## Data Availability

The trial data used for the cumulative analysis is available from the cited publications. We share the complete Trial Sequential Analysis files alongside the publication.

## Acknowledgements

Copenhagen Trial Unit for providing salaries to the authors.

## Appendix

**Figure 1.**
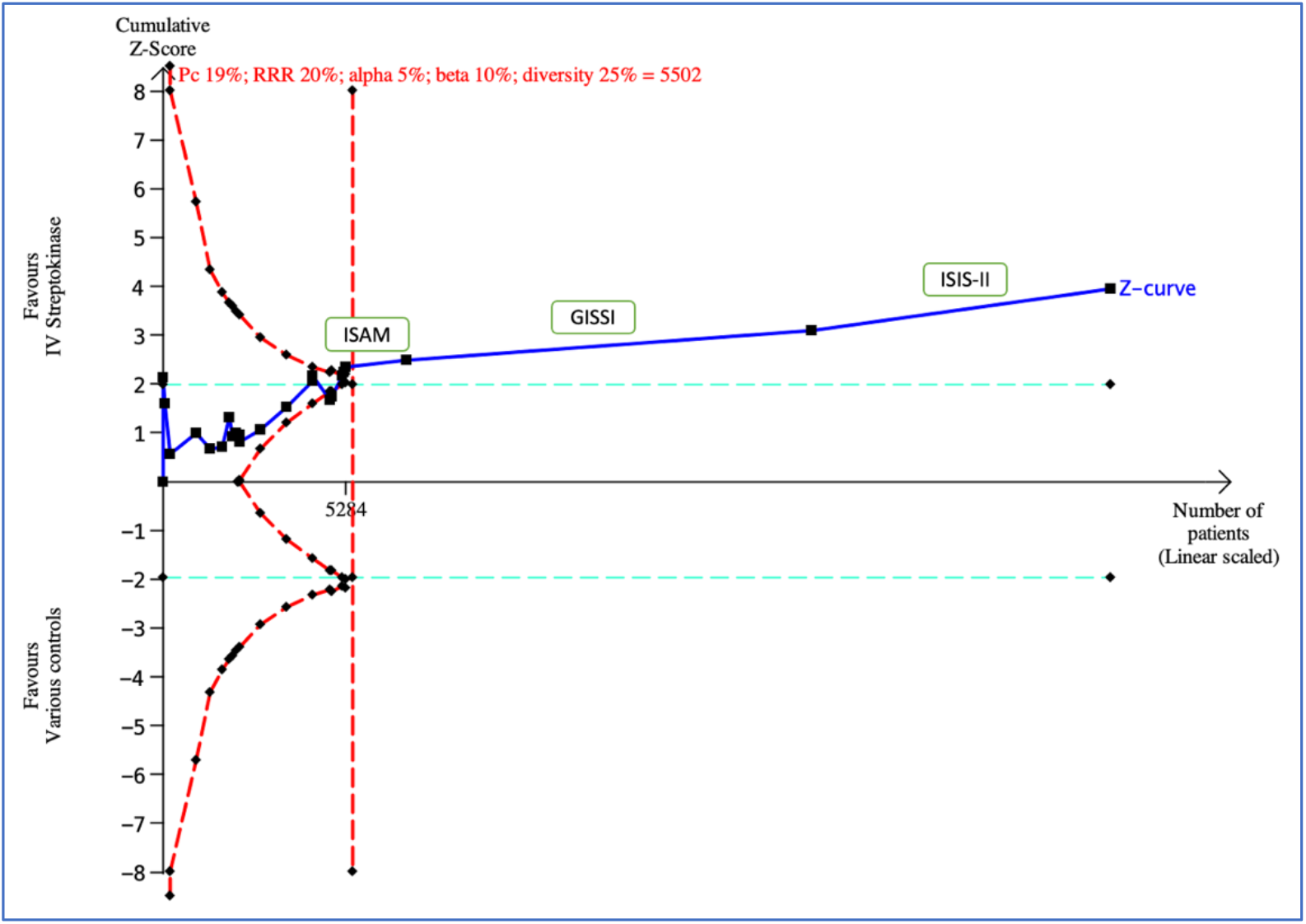
Sensitivity analysis 1: Diversity=25%.

**Figure 2.**
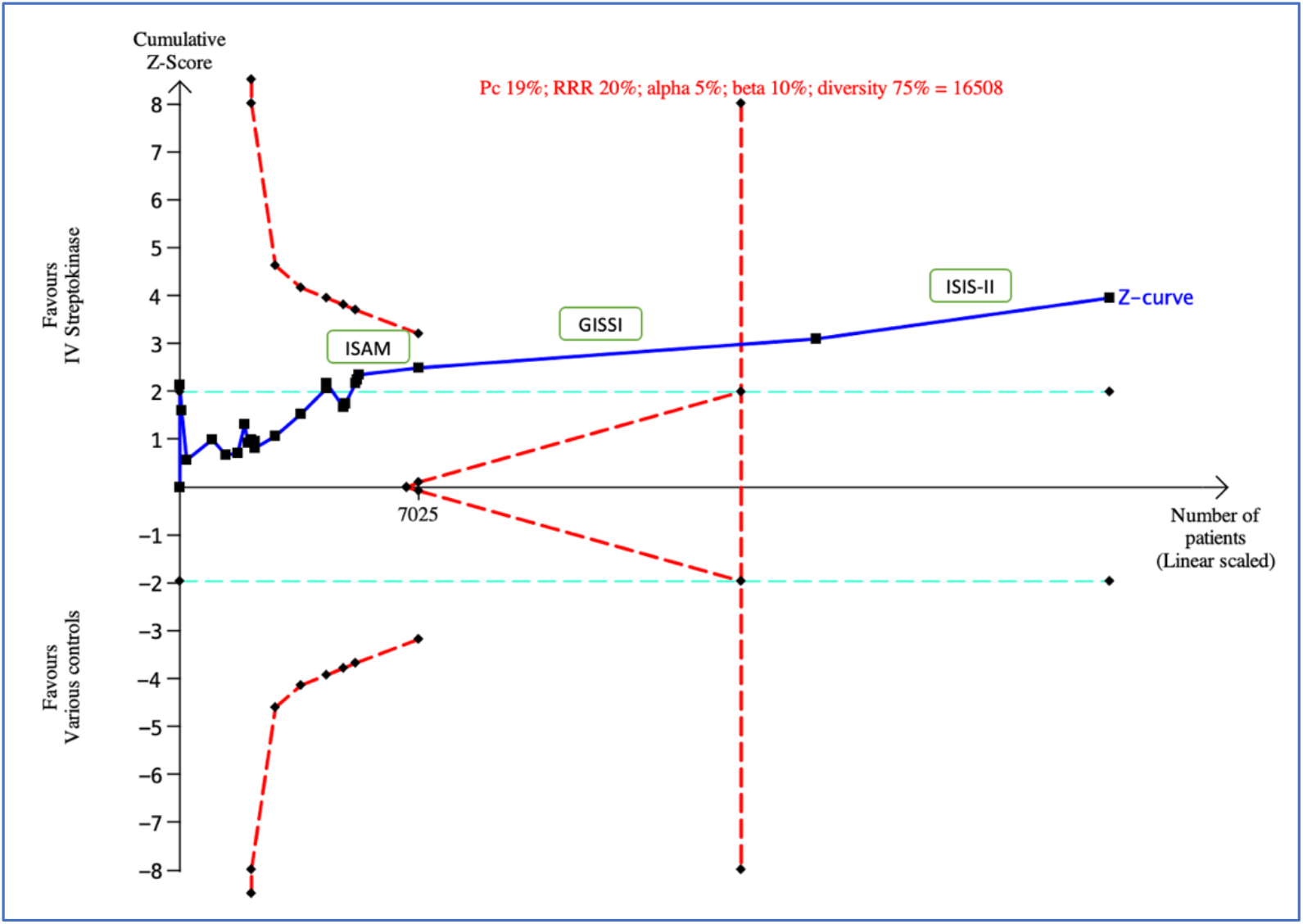
Sensitivity analysis 2: Diversity=75%.

**Figure 3.**
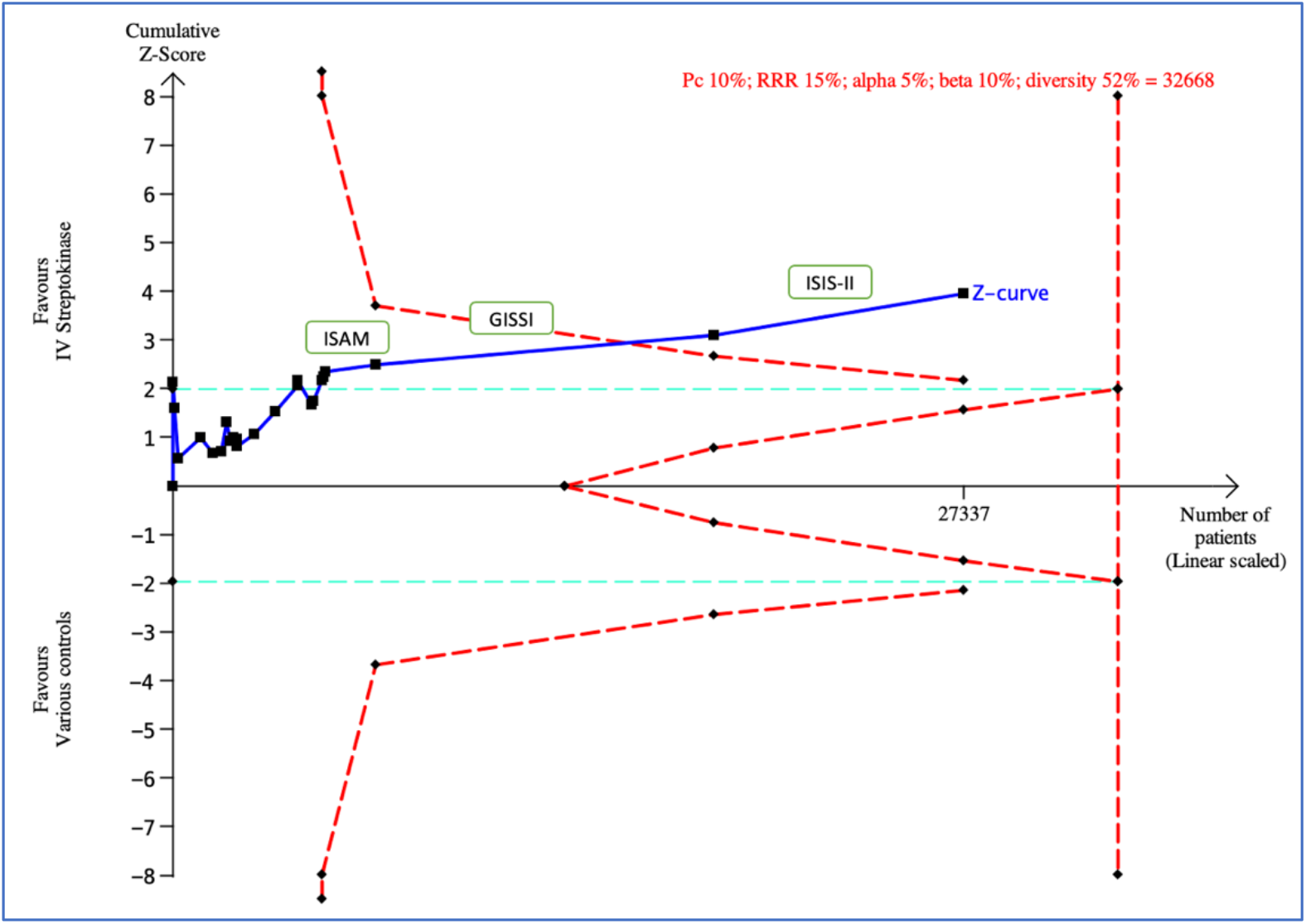
Sensitivity analysis 3: based on Pogue and Yusuf’s assumptions of parameters. Pogue and Yusuf assumptions (control event proportion 10%, relative risk reduction (RRR) 15%, alpha 5%, beta 10% (power 90%))^6,7^ plus diversity 52%.

**Figure 4.**
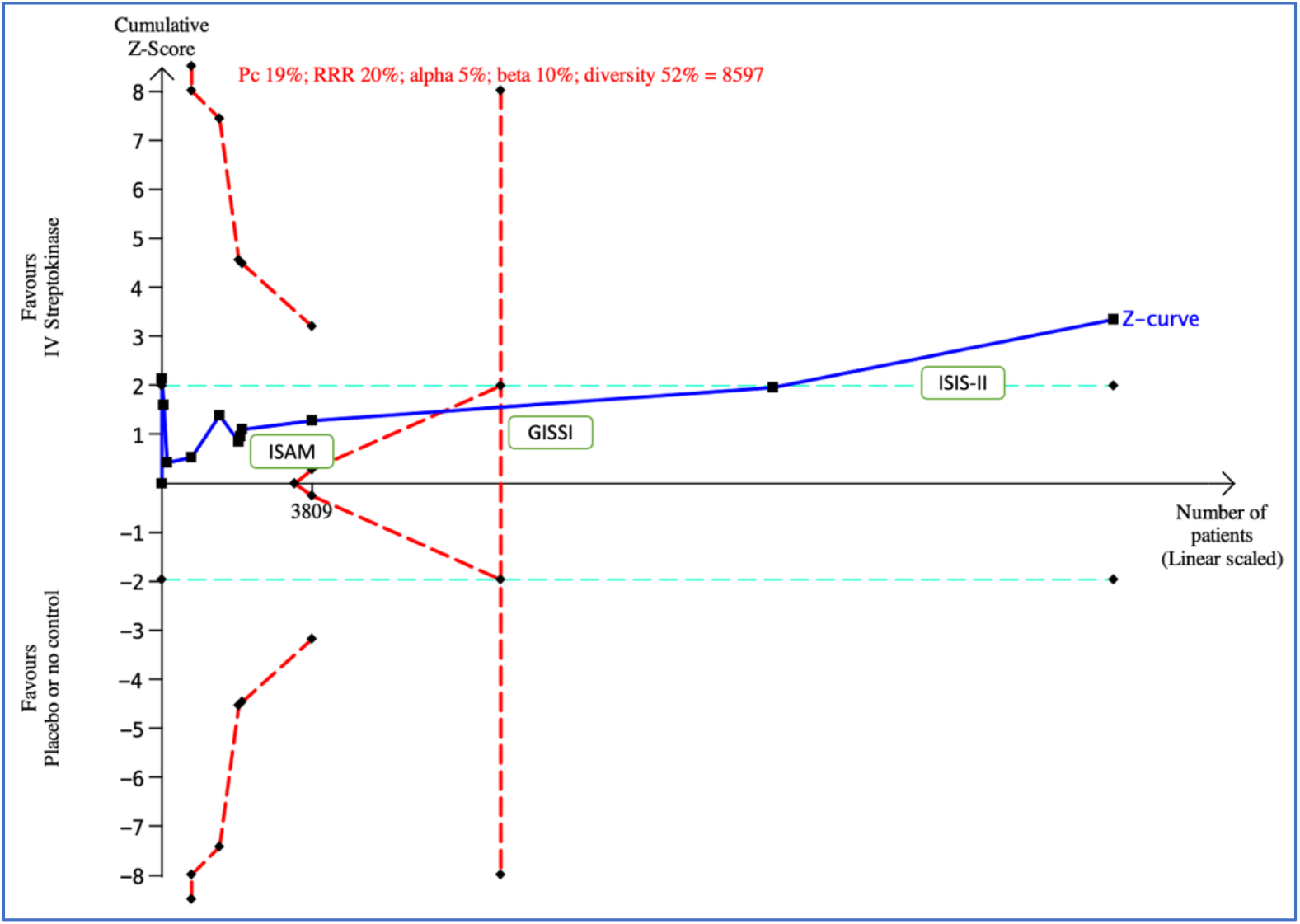
Sensitivity analysis 4: Placebo-controlled or no intervention-controlled trials only. Including placebo-controlled (Fletcher 1959, Dewar 1963, Olson 1984, Schreiber 1984, ISAM, ISIS-II) and no intervention controlled (Austrian 1977, NHLBI 1974, UK Collab 1976, N German Collab 1977, GISSI) trials.

